# Socioeconomic determinants of mobility responses during the first wave of COVID-19 in Italy: from provinces to neighbourhoods

**DOI:** 10.1101/2020.11.16.20232413

**Authors:** Laetitia Gauvin, Paolo Bajardi, Emanuele Pepe, Brennan Lake, Filippo Privitera, Michele Tizzoni

**Author notes:** these authors contributed equally to this work (Laetitia Gauvin, Paolo Bajardi).

## Abstract

As the second wave of SARS-CoV-2 infections is surging across Europe, it is crucial to identify the drivers of mobility responses to mitigation efforts during different restriction regimes, for planning interventions that are both economically and socially sustainable while effective in controlling the outbreak. Here, using anonymous and privacy enhanced cell phone data from Italy, we investigate the determinants of spatial variations of reductions in mobility and co-location in response to the adoption and the lift of restrictions, considering both provinces and city neighbourhoods. In large urban areas, our analysis uncovers the desertification of historic city centers, which persisted after the end of the lockdown. At the province level, the local structure of the labour market mainly explained the variations in mobility responses, together with other demographic factors, such as population’s age and sex composition. In the future, targeted interventions should take into account how the ability to comply with restrictions varies across geographic areas and socio-demographic groups.

## 1 Introduction

In the effort of fighting the COVID-19 pandemic, several governments worldwide have imposed unprecedented mobility restrictions and social distancing policies, as these - combined with contact tracing and isolation of cases - represent the most effective strategy to slow down the spread of SARS-CoV-2 [1, 2, 3, 4, 5] in absence of a vaccine.

Since the very beginning of the COVID-19 pandemic, changes in human mobility ensuing the non-pharmaceutical interventions (NPIs) adopted in many countries worldwide have been measured through the analysis of mobile phone data [6]. To mention a few examples, previous studies have investigated changes in human movements through mobile phone data in Austria, China, Japan, the UK, Germany, and the United States [7, 8, 9, 10]. Several of these studies suggested that mobility restrictions unevenly impact different socioeconomic strata and that income inequalities are associated to a different capacity to afford prolonged social distancing [11, 12, 13].

While it has been observed that income disparities are strongly associated with differences in mobility reductions during a lockdown, the responses to milder mitigation policies, and how such policies may impact different areas of a country depending on their local economies, have not been explored in detail. In particular, the study of mitigation policies in metropolitan areas and their impact in different neighbourhoods, remains limited to a few paradigmatic examples, like New York City [8]. As Europe is facing a rapid resurgence of COVID-19 cases, new measures to reduce transmission will be needed for some time and tailored solutions as an alternative to a blanket lockdown need to be developed. To understand which policies might be both sustainable and effective in controlling the outbreak, it is crucial to identify the drivers of mobility responses both during the lockdown and when restrictions are gradually lifted.

Here, we extensively investigate the socioeconomic determinants of the responses to mobility restrictions imposed in Italy during the full course of the first wave of the COVID-19 epidemic, from February til June 2020, through the analysis of human mobility patterns derived from anonymized and aggregated mobile phone data. To this aim, we gauged the mobility responses during 3 phases of the spring COVID-19 wave in Italy: first, at the beginning of the outbreak, before the enforcement of the national lockdown; second, during the lockdown, and, third, immediately after the lift of the lockdown. We mapped mobility changes onto different spatial scales, at level of the Italian provinces and at a finer granularity, namely that of city districts in three major metropolitan areas: Turin, Milan and Rome. Mobility changes induced by self-initiated behavioral responses and by top-down interventions displayed a significant heterogeneity across all spatial scales. To investigate the socioeconomic determinants that may have driven such geographic variations, we modeled their relationship with a number of demographic, economic, and epidemiological covariates, including - among others - the fraction of workers by economic sector, the average personal income, the fraction of women in the population, the number of commuters and the timing of interventions. The results show that behavioral responses were associated to several socioeconomic factors, including but not limited to income inequalities, in the different phases of the pandemic management cycle and across different geographic scales. Our findings shed a light on the complex landscape of the determinants of responses to the introduction of NPIs and their relaxation.

## 2 Results

### 2.1 Spatial variations of responses to social distancing orders in Italy

To quantify the mobility responses to NPIs across Italy, we computed two mobility metrics: the radius of gyration *r*_*g*_ and the mean degree of the co-location network ⟨*k*⟩. Such metrics were evaluated on a daily basis for each of the 107 Italian provinces. Both metrics are related to users’ mobility but they capture different complementary aspects of it. The radius of gyration is averaged over all users who live in a given province (see Methods for more details on home assignment), and it provides a measure of the spatial range covered by their daily movements. The co-location network is built considering all the users present in a given province every hour, and therefore it takes into account the co-presence of people who may live far away from each other.

Both mobility metrics displayed a sharp decline with respect to their baseline values immediately after the first official report of a cluster of COVID-19 cases in Lombardy, on February 21, 2020 (Fig. 1). Initially, such decline was mainly due to self-induced behavioral changes, since mobility restrictions and stay-at-home orders were imposed only onto a few relatively small areas of Northern Italy. As social distancing orders became tighter, *r*_*g*_ and ⟨*k*⟩ continued to drop until a national lockdown was declared on March 11. During the lockdown, both metrics plateaued at an average −70% relative reduction with respect to the baseline, with distinct weekly patterns due to the movements of the active workforce employed in essential services. On May 4, the lockdown was lifted, the so-called phase 2 started and the mobility trends reversed, although without reaching the baseline values, even after 3 weeks since the reopening.

**Figure 1:**
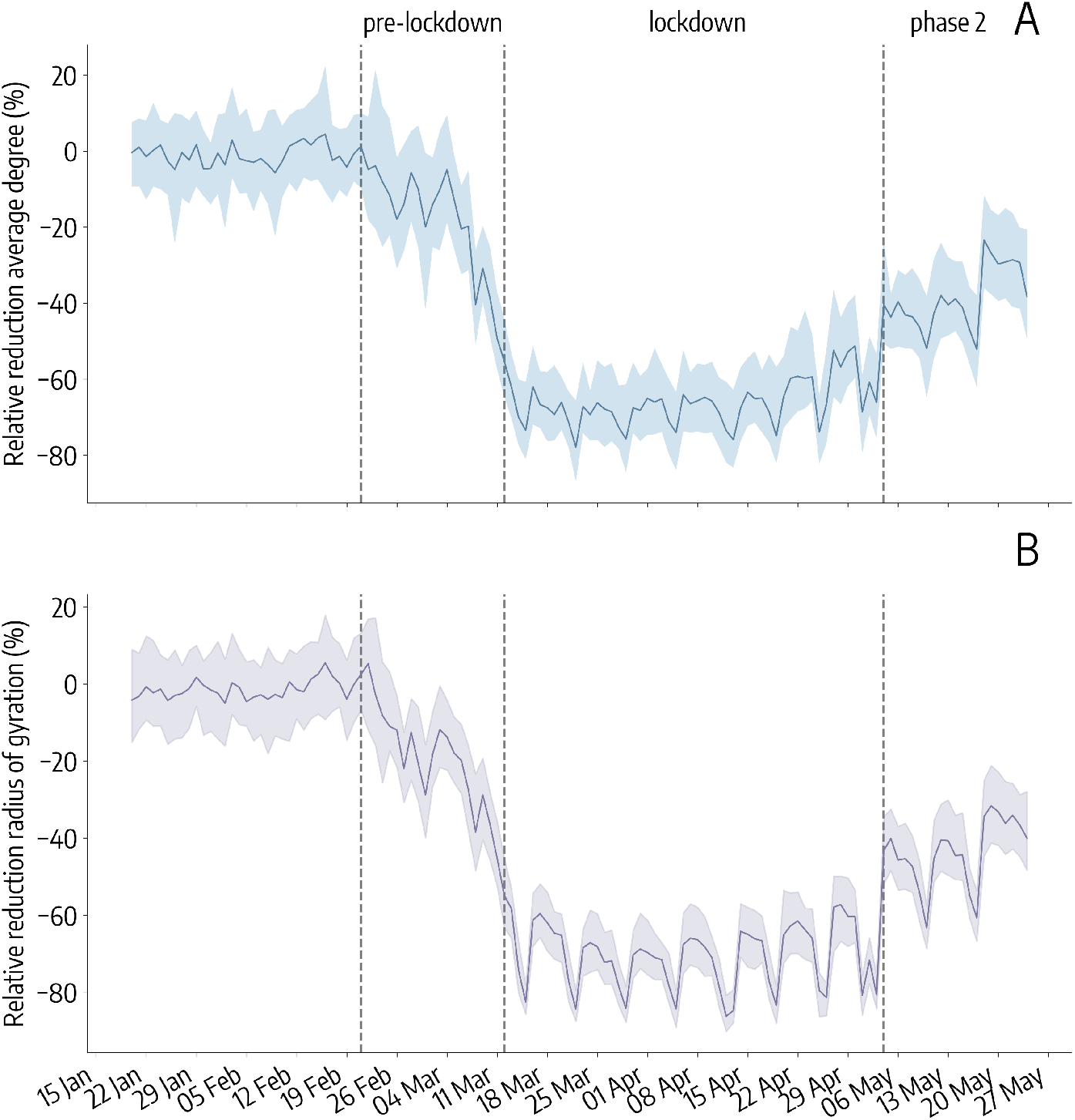
Mobility responses during the COVID-19 pandemic in Italy. Time series of the relative reduction with respect to the baseline of the average degree of the co-location network (A) and the median radius of gyration (B) in Italy. The solid line indicates the median value computed across the 107 Italian provinces, while the shaded area corresponds to the 50% reference range. Dashed vertical lines mark the dates that define the 3 phases of the pandemic response: the Pre-lockdown (between February 21 and March 11), the Lockdown (between March 12 and May 4) and the Phase 2 (after May 4).

Although the general mobility trend was consistent at the national level, the relative reductions of *r*_*g*_ and ⟨*k*⟩ displayed a high spatial heterogeneity. A map of the relative reductions of *r*_*g*_ (Fig. 2) and ⟨*k*⟩ (Fig. S1) shows that users’ mobility did not decrease uniformly across provinces during the pandemic period. For instance, on March 6 (Fig. 2 left panel), before the lockdown, the median *r*_*g*_ had decreased by 44% in Lodi and Rimini, with respect to the baseline levels, but it had actually increased by 9% in Aosta and by 25% in Bolzano. During the lockdown, the reduction in mobility was more uniform across the country, yet showing variations between provinces. On March 20, the median *r*_*g*_ had decreased by 80% in Rome and by 53% in Bolzano. Finally, at the start of phase 2 some provinces returned more quickly to the baseline mobility levels, while others maintained higher levels of social distancing. On May 18, when all economic activities reopened, the median *r*_*g*_ had reached baseline levels in Aosta (+2%) and Sondrio (+2%) but remained at -47% with respect to the baseline in Milan.

**Figure 2:**
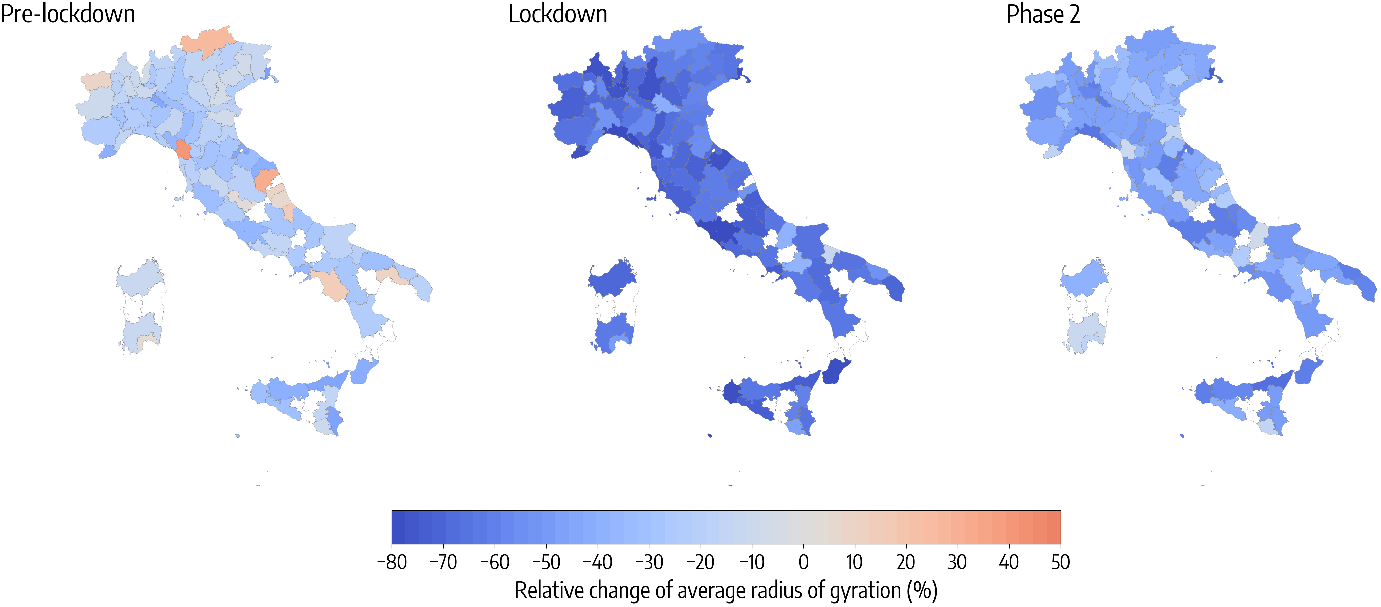
Spatial patterns of mobility reductions. Each map shows the relative change of the median radius of gyration with respect to the baseline, by province, on 3 different days, one for each period under study: the Pre-lockdown (left), the Lockdown (center) and the Phase 2 (right). Provinces in white are not included in the analysis because of low population sampling (less than 100 users).

### 2.2 Socioeconomic determinants of mobility responses across the public health intervention cycle

At the level of Italian provinces, we examined the association between the reductions in mobility and several socioeconomic and epidemiological features through a multivariate regression analysis with variable selection (see Methods for more details). The importance of the association of each covariate with the mobility changes can thus be quantified through the corresponding regression coefficient *β* and its 95% confidence interval. Fig. 3 shows the variables that we found to be associated with the daily reductions in the radius of gyration, in 3 weeks corresponding to the 3 phases of the first pandemic wave: the pre-lockdown (March 2-8), the lockdown (March 16-22) and the phase 2 (May 11-17). Here, only three weeks are shown, but the results remain consistent along the full period of the first wave (see Fig. S4). The results of the same analysis for the reductions of ⟨*k*⟩ are reported in Fig. S3, and the values of the regression coefficients and their confidence intervals in all the 3 weeks for the two mobility metrics are shown in Tab. S1-S6.

**Figure 3:**
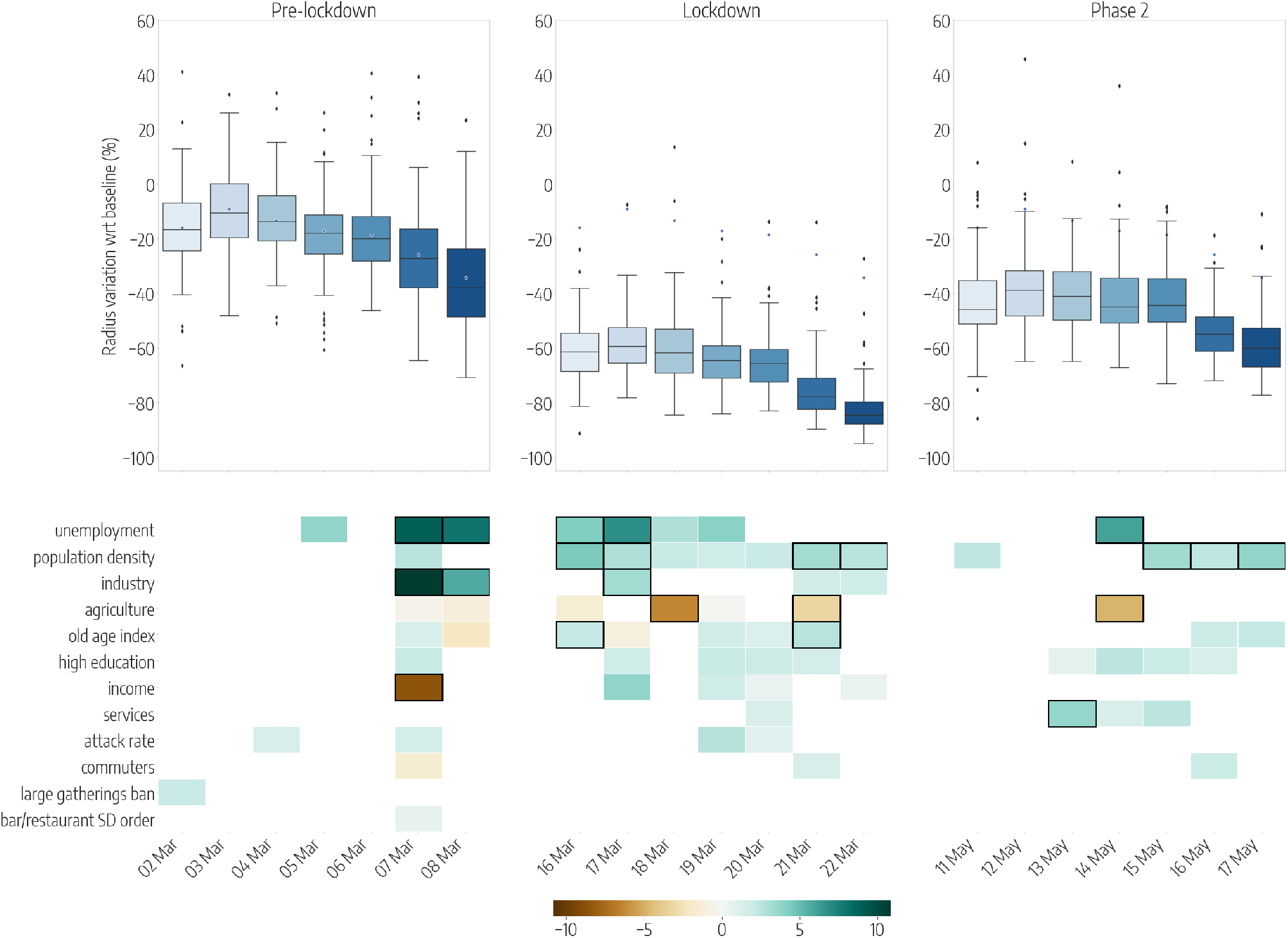
Province-level mobility reductions and their socioeconomic determinants. Top panels show the relative reductions in the median radius of gyration with respect to the baseline, each day (from Monday to Sunday) in 3 different weeks of the periods under study: the Pre-lockdown (left), the Lockdown (center) and the Phase 2 (right). Boxplots describe the distributions of the reduction by province. Bottom panels show the results of the regression model, by week, displaying the socioeconomic variables that are selected by the model to predict the corresponding variations in mobility, on each day. Coloured boxes correspond to those covariates that are selected by the model, and the color codes the associated coefficient in the regression, from negative (brown) to positive (blue). Black solid squares highlight the covariates that are statistically significant at *p <* 0.05.

In general, economic variables, and in particular those related to the distribution of the workforce across labour sectors, were most frequently selected by the model as significantly associated to the mobility responses. Workers in the industrial and the service sectors were those most affected by the restrictions that closed all non-essential businesses, and they were encouraged to work from home once the restrictions were lifted. Indeed, provinces with a larger share of their workers employed in the industry sector experienced a larger reduction of *r*_*g*_ before (*β* = 10.9, 95% CI [5.7, 16.0] on March 7) and during the lockdown (*β* = 3.2, [0.6, 5.9] on March 17). Similarly, a higher proportion of workers in the services was associated to a higher reduction of mobility after the lift of the lockdown (*β* = 3.7, [0.8, 6, 5] on May 13). On the other hand, provinces characterized by a larger agricultural workforce were characterized by a relative smaller reduction of *r*_*g*_, both during the lockdown and in phase 2 (*β* = −6.4, [−9.5, *−*3.2] on March 18, and *β* = −4.8, [−8.4, *−*1.2] on May 14). This can be expected as the entire agricultural sector remained fully active to ensure food provisioning, even during the strictest lockdown period. Higher levels of unemployment were associated to a larger reduction of *r*_*g*_ in all the 3 phases: before the lockdown (*β* = 9.1, [1.8, 16.4] on March 7), during the lockdown (*β* = 6.9, [2.8, 10.9] on March 17) and after the ease of the restrictions (6.0, [1.9, 10.1] on May 14). Income was generally positively associated - although not significantly - with larger mobility reductions during the lockdown (*β* = 3.8, [−0.1, 7.7] on March 17). Interestingly, a higher income was found to predict an increase in mobility (*β* = −8.5, [−16.1, *−*0.9]) on a single day, March 7, that was the Saturday before a national stay-at-home order took effect. This result can be explained by the sudden relocation of people, out-of-home workers and students, from large cities to their place of origin, in urban belts, that followed the rumors of an imminent lockdown [14].

From a demographic perspective, we found that an older and a more educated population was positively associated with larger mobility reductions during the lockdown and in phase 2. The regression coefficients for the old-age index were *β* = 2.2, [0.0, 4.5] on May 16, and *β* = 2.7, [0.7, 4.7] on March 21. Overall, higher education levels were positively associated to the reductions across the 3 phases even though they were not statistically significant.

Population density was frequently associated with larger mobility reductions, both during the lockdown and after (*β* = 4.4, [2.2, 6.6] on March 16, *β* = 3.4, [0.7, 6.1] on May 15). The effect of population density is particularly evident during weekends (March 21-22 and May 16-17) as mobility in densely populated urban areas was highly limited, while long range travel to reach essential services was unavoidable in rural areas.

Finally, since the spread of SARS-CoV-2 in Italy was highly heterogeneous and in some provinces NPIs were implemented earlier than in others, we controlled for the epidemic activity and the adoption of NPIs by including a number of epidemiological variables in our analysis. Overall, the attack rate, that is the cumulative fraction of reported infections in a province on a given date, was mildly positively associated - yet not significantly - with larger reductions of *r*_*g*_. Also, large gathering bans and closures of bars and restaurants did not show a significant effect on *r*_*g*_. However, social distancing policies had a much larger effect in reducing the average degree of the co-location network, ⟨*k*⟩, before the lockdown (see Tab. S4). For instance, the ban of large gatherings was associated to a larger reduction in ⟨*k*⟩ on March 4 (*β* = 8.4, [1.4, 15.3]) together with a similar effect of bar and restaurant closures on March 3 (*β* = 6.2, [2.0, 10.4]), suggesting these measures were effective at the time of adoption. Larger reductions of ⟨*k*⟩ were also associated with a higher income in every phase of the outbreak (*β* = 6.3, [2.4, 10.2] on March 5, *β* = 2.9, [0.0, 5.7] on March 20, and *β* = 3.3, [−0.1, 6.7] on May 13), and overall, the determinants of reductions in ⟨*k*⟩ were consistent with those observed in the case of *r*_*g*_, but providing in general lower values of r-squared with respect to the model of the radius of gyration.

### 2.3 Mobility responses in the neighbourhoods of large metropolitan areas

By looking at mobility changes in urban areas, we similarly observed a high spatial variability of responses across the districts of three main Italian cities: Turin, Milan and Rome. In Fig. 4 the reduction of the average degree of the co-location network, ⟨*k*⟩ in the 3 cities is shown for three days during the pre-lockdown, the lockdown and the phase 2 (corresponding to the same dates of Fig. 2). Turin and Milan, in the North, experienced an early decline in ⟨*k*⟩ before the lockdown and even in absence of targeted measures, thus hinting to the presence of self-initiated behavioral changes. Instead, on March 6, the value of ⟨*k*⟩ had decreased with respect to the baseline only in some districts of Rome but not in others. Differences in social distancing lowered during the lockdown, and all cities experienced a deep reduction of ⟨*k*⟩, ranging between −30% and −80%, depending on the district. Overall, all the cities share a common pattern: the most central districts were those displaying the largest reduction of ⟨*k*⟩, since the start of the outbreak until the ease of the restrictions. Once restrictions were lifted, mobility increased more quickly in the periphery, leaving the historic city centers empty for a longer time period.

With respect to the province level, the different geographic scale of the urban areas prompted us to focus on the co-location network as the most suitable metric to measure mobility responses. The higher spatial granularity allows to better capture differences in ⟨*k*⟩, while estimates of *r*_*g*_ are affected by the small sample size of users who are assigned to each district. As reported in Fig. 5, the most important feature that was associated to a larger reduction in the average degree of the co-location network, during the whole outbreak, is the proportion of people who completed a higher education degree (*β* = 20, [1.7, 38.2] on March 6, and *β* = 12.7, [4.7, 20.7]) on March 18). This result is intuitive, as neighbourhoods characterized by higher education levels are also the most affluent and those with a larger fraction of residents employed in professional services.

**Figure 4:**
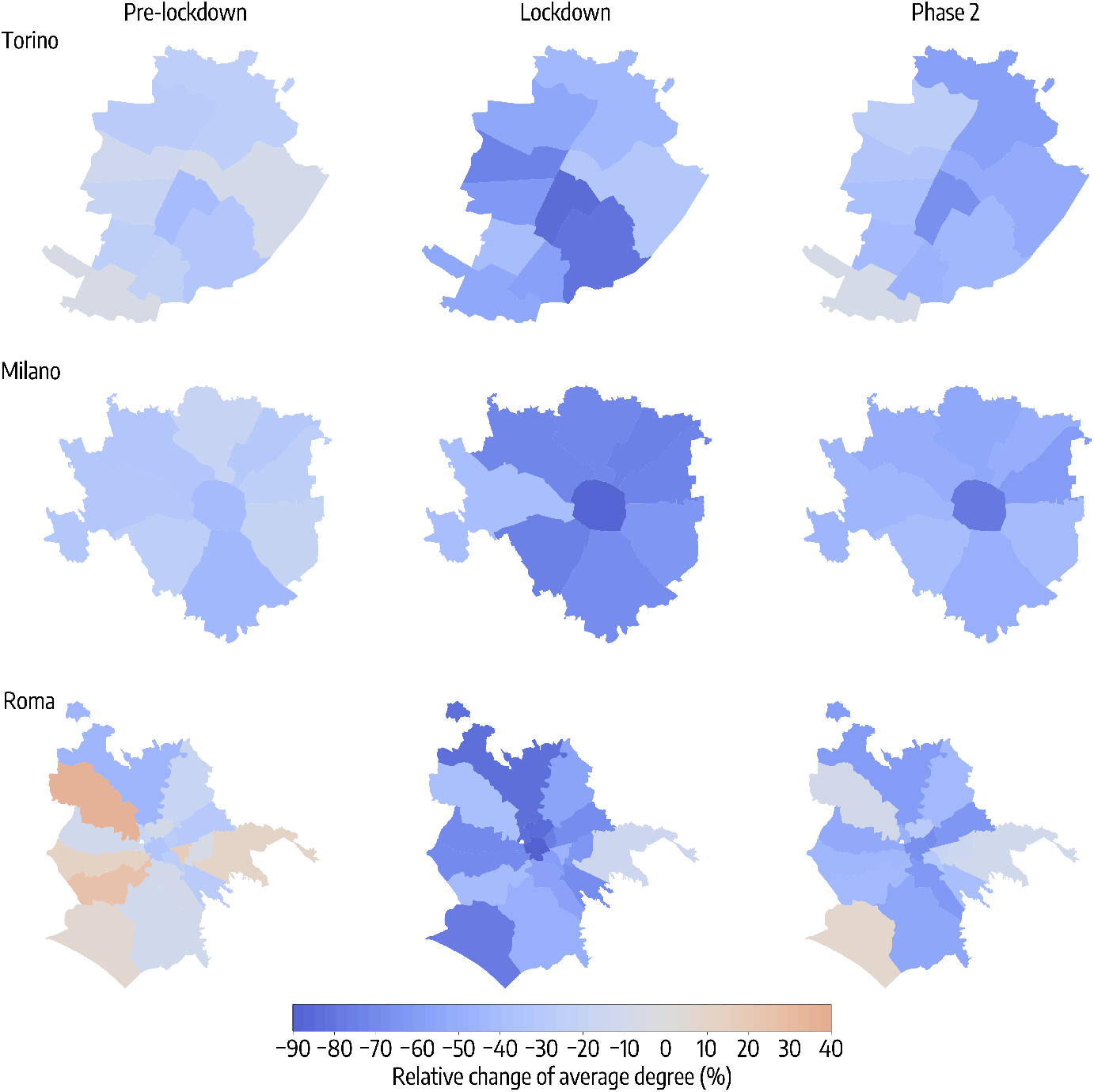
Spatial patterns of variations in co-location in metropolitan areas. Maps of the relative reduction of the average degree of the co-location network in Turin (top), Milan (middle) and Rome (bottom), by city district, on 3 different days, one for each period under study: the Pre-lockdown (left), the Lockdown (center) and the Phase 2 (right).

**Figure 5:**
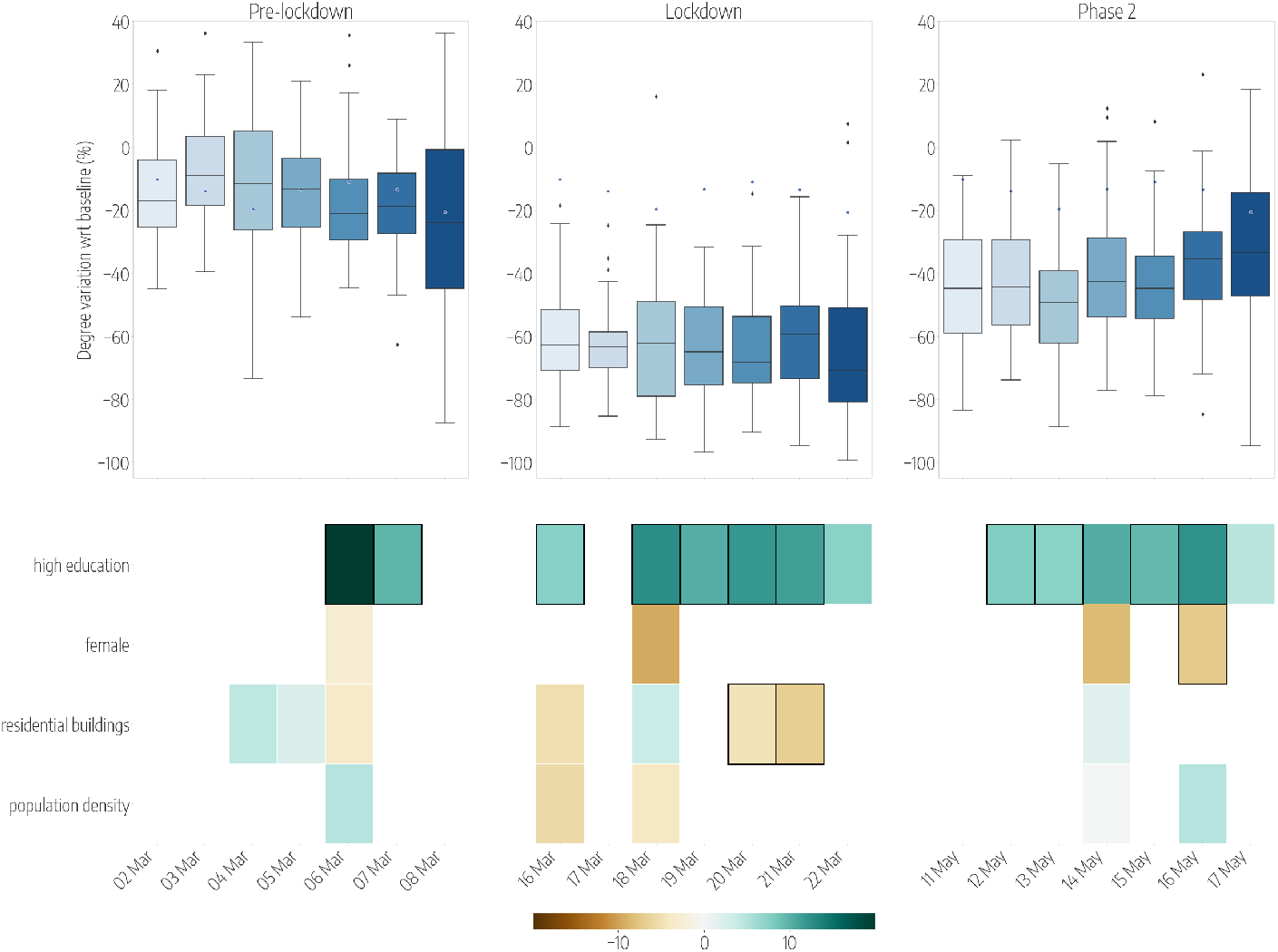
Predicted district-level variations of spatial co-location and their socioeconomic determinants. Top panels show the relative reductions in the median degree of the co-location network with respect to the baseline, each day (from Monday to Sunday) in 3 different weeks of the periods under study: the Pre-lockdown (left), the Lockdown (center) and the Phase 2 (right). Boxplots describe the distributions of the reduction by city district, considering all the districts of Turin, Milan and Rome. Bottom panels show the results of the regression model, by week, displaying the socioeconomic variables that are selected by the model to predict the corresponding variations in co-location, on each day. Coloured boxes correspond to those covariates that are selected by the model, and the color codes the associated coefficient in the regression, from negative (brown) to positive (blue). Black solid squares highlight the covariates that are statistically significant at *p <* 0.05.

Among the other features considered, a higher proportion of women appears to be negatively associated with the reduction of ⟨*k*⟩. On May 16, the percentage of female appears to be statistically significant with a negative coefficient *β* = −7.1, [−14.2, *−*0.1], thus indicating a higher level of co-location in places with a higher proportion of female residents. After adjusting for population density, which is never significant at this spatial resolution, a larger share of residential buildings (i.e. buildings that are not used as offices, shops or for manufacturing activities) was associated with a smaller relative reduction in ⟨*k*⟩ during the lockdown.

## 3 Discussion

The behavioral responses to the mitigation policies adopted by the Italian government during the first wave of the pandemic differed substantially across Italy. The spatial heterogeneity in mobility responses cannot be explained by the geographic differences observed in the spread of SARS-CoV-2, only.

Mobility reductions were observed to significantly vary both at the province level and at the city-district level. In particular, in large urban areas, our data-driven analysis exposed an interesting phenomenon with several social and economic consequences: the desertification of historic city centers. As many urban centers worldwide have been severely hit by the pandemic, public health interventions prompted the relocation of residents from large cities to the countryside, as observed, for instance, in France [15] and in the USA [16]. Our analysis shows the presence of such effect in three major Italian cities, before and after the end of the lockdown, thus leading to a persistent divide in mobility between the center and the periphery.

According to our study, the geographical differences are mainly explained by the economic factors related to the local structure of the labour force, with different phases and their related mitigation policies impacting different category of workers. Before and during the lockdown, mobility decreased more in provinces with a higher proportion of employees in the industry sector, as many of them were laid off with wage compensations. It also decreased more in the presence of higher unemployment, suggesting the existence of an additional burden of the restrictions in those areas characterized by low employment levels. The share of employees in the service sector did not explain much of the variation in mobility changes before the lockdown, but it became an important driver during the reopening as it became positively associated to mobility reductions. A possible explanation is that a large proportion of people working in services were either still working remotely or still not allowed to work [17]. Indeed shops, bars, restaurants and department stores were still closed on the second week of May. Moreover remote working was widely implemented as much as possible since the early start of the outbreak and individuals whose occupation was more suitable for remote working maintained their arrangements after the end of the lockdown. Our results are in line with early observations about the disproportionate impact of the interventions due to income inequalities [18, 19], and of recent studies on the effects of restrictions on local labour market areas [20], but expand such findings by highlighting the importance of the structure of the labour force.

While most of the variations in mobility are linked to the structure of the workforce share, demographic factors still played a role in the heterogeneity of the mobility responses, even though to a lesser extent. Across provinces, a higher ratio of elderly to young population was associated with a higher reduction in mobility. Previous survey-based studies have shown that non-adherence to mitigation policies in the UK was associated with a younger population, lower socioeconomic grade and working in a key sector [21]. In urban areas instead, we found a larger female population to be associated with smaller mobility reductions, in contrast to previous survey studies [22] and to previous research investigating the stay-at-home compliance during the H1N1 2009 pandemic [23]. In this case, our result may suggest responses to COVID-19 in Italy were disproportionately affecting population strata that might be more prone to comply with restrictions but could not, because financially distressed. Finally, while the outbreak were highly clustered in its early phase, we did not observed a strong effect of risk perception (measured by province specific attack rates) in driving the mobility reductions in Italy, as it was observed in France [15].

We acknowledge some limitations in our study. While the distribution of users resemble fairly well the distribution of the actual population both at the province level and at the district level within cities, we expect our sample to be skewed towards better educated, wealthier and younger users [24]. Moreover, anonymous digital traces do not allow to investigate and adjust for personal beliefs and people’s intrinsic motivation, including substantive moral support and social norms and expectations about how long the measures would be in place, that are known to affect intentions to comply [25, 26]. Finally, our study being of an observational type, caution is needed before extrapolating a direct causal effects between covariates and the observed changes in human behaviour.

In conclusion, our work underscores the important role of socioeconomic factors and in particular of the labour structure [27] in shaping behavioral responses during the full course of the pandemic cycle, from early interventions to the reopening. In particular, our approach highlighted the unequal impact of mobility restrictions in urban areas, where central districts experienced a much more prolonged reduction of mobility and social contacts than the periphery. This has policy implications for the management of the pandemic in many cities worldwide, especially those characterized by large socioeconomic inequalities [28]. Future intervention policies to mitigate the epidemic and hamper the economic shock, should take into account the extent to which specific population strata are able to comply. Horizontal restrictions might work in theory but will induce different local responses in practice because of the underlying social differences. Besides mobility limitations, adequate economic incentives [29, 30] and behavioral nudging [31, 32] should be implemented to maintain high levels of compliance, while additional strategies should be devised and implemented locally to protect individuals who can not afford to comply with mobility restrictions [33, 34, 35].

## 4 Methods

### 4.1 Location Data

We analyze location data provided by Cuebiq Inc. Location is collected anonymously from opted-in users, who provided access to their location through a GDPR-compliant framework. The operating system of the device (iOS or Android) combines various location data sources (e.g. GPS, WiFi networks, mobile network, beacons) to provide geographical coordinates together with an estimate of measure accuracy. Several factors may affect location accuracy (that can also vary over time for the same device), but it can be as accurate as 10 meters. The data and the mobility metrics are extensively described elsewhere [36]. Here, we recall that the basic unit of information we process is an event of the form (`anonymous hashed user id, time, latitude, longitude`), which we call a user’s *stop* in the remainder. We perform all our analyses on a panel of about 41,000 anonymous, opted-in users for whom there was at least one stop collected during every week from January 20 to May 24. This led to about 300 million data points over the 18 weeks of this study. We average different mobility and proximity metrics during the pre-outbreak period (between January 20 and February 21, 2020) and observe their weekly and daily evolution over the course of the outbreak.

### 4.2 Definition of home location

We assign each user to a province of residence (“home location”) considering only her traces in the pre-outbreak period. We assume that home location is the most frequently visited night time location [37]. Thus we define the home province to be the province where a user has spent most of the time within the time interval 00:00 -6:00, between 18 January and 21 February 2020. We consider all the stops whose duration has an intersection with the interval 00:00–06:00. For the users with home location in the provinces of Turin, Milan and Rome we further assign them to one of the city districts (or having home out of the urban area) with the same approach.

Once all users have been assigned to a home province or district, we compared their distribution with respect to official census statistics. Our users’ sample matches fairly well the distribution of the Italian population across provinces (see Fig.S1) and in the districts of the three cities under study (see Fig.S2).

### 4.3 Socioeconomic and epidemiological covariates

Candidate covariates are taken from official data sources, as reported in Tab. 1: the Italian National Institute of Statistics (ISTAT), the Ministry of Economy and Finance (MEF) and the Civil Protection Department (CPD) in charge of coordinating the national surveillance of SARS-CoV-2 (http://github.com/pcm-dpc/COVID-19). Some variables are available only at the aggregated level of provinces (e.g. number of employee of a certain productive sector), while others are collected at the finest spatial resolution of census block (e.g. number of commuters) and thus included in the analysis of city districts.

Epidemic variables are not included in the analysis of city districts because restrictions were not differentiated by neighbourhoods and because SARS-CoV-2 incidence data is not available at that spatial resolution. Since we are interested in investigating the local heterogeneities in behavioural changes at the district level, every variable has been normalized at the city level and only city-specific z-scores for every variable have been used in the model. In such a way, we were able to measure the effect of the different covariates, disregarding the baseline values of the three cities.

### 4.4 Mobility metrics and mobility changes

To assess the effect of NPIs on our users’ sample, we compute two metrics that capture different notions of mobility. An extensive description of the data processing workflow to generate these metrics is reported in [36].

The first metric is an individual feature, the radius of of gyration of a user, *r*_*g*_, a common measure of the spatial range covered by a user’s mobility patterns [38]. It is defined as:

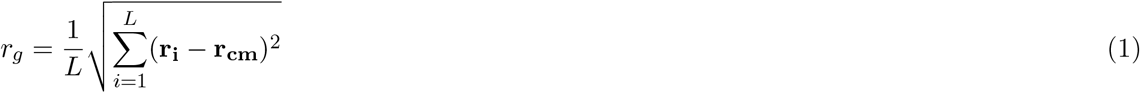

where *L* is the full set of stops made by a user over a given time frame, **r**_**i**_ is the vector of coordinates of stop *i* and **r**_**cm**_ is the vector of coordinates of the center of mass, weighted by the duration of each stop Δ*t*_*i*_. We compute the radius of gyration for each user on a daily basis, so that *L* in Eq. 1 represents the set of stops made by a user during a day.

The second metric is the average degree of a spatial co-location network that is a proxy of potential social mixing of the population. To build the network we collect all the positions of all users in a given province within time windows of 1 hour, and we connect any two individuals with an edge if they made at least one stop within a distance *d* = 100 m from one other, within the same hour of the day. We then compute the mean hourly network degree ⟨*k*⟩ = 2*E/N*, where *E* is the number of edges and *N* is the total number of nodes in the network, including those with *k* = 0. By taking the average of ⟨*k*⟩ over all the 1-hour slices of a given day, we obtain a daily average degree for every province.

**Table 1:** Description and data source of the explanatory variables.

| Variable | Source | Province/<br>Urban | Year | Description |
| --- | --- | --- | --- | --- |
| **Demography** |  |  |  |  |
| Old age index | ISTAT | P/U | 2011 | Ratio of people >65 years over people <15 years old |
| Females | ISTAT | U | 2011 | Percentage of females |
| Higher Education | ISTAT | P/U | 2011 | Percentage of people with high school degree or higher education degree (as highest certification) |
| **Economy** |  |  |  |  |
| Income | MEF | P | 2018 | average income (kEuro) |
| Unemployment | ISTAT | P/U | 2011 | Percentage of unemployed labour force |
| Commuters | ISTAT | P/U | 2011 | Percentage of people commuting daily outside the municipality of residence |
| Agriculture | ISTAT | P | 2019 | Percentage of employees in the Agriculture sector |
| Industry | ISTAT | P | 2019 | Percentage of employees in the Industry, Manufacturing and Constructions sector |
| Services | ISTAT | P | 2019 | Percentage of employees in the Services, Retail, Tourism sector |
| **Territory** |  |  |  |  |
| Population density | ISTAT | P/U | 2017 | Population per unit area |
| Residential buildings | ISTAT | U | 2011 | Percentage of buildings for dwelling purposes |
| **Epidemic variables** |  |  |  |  |
| Attack rate | CPD | P | 2020 | Percentage of COVID-19 positive tests |
| Bar/Restaurant SD order | CPD | P | 2020 | Early phase mitigation policy (binary variable) |
| Large gathering ban | CPD | P | 2020 | Early phase mitigation policy (binary variable) |
| Mandatory school closure | CPD | P | 2020 | Early phase mitigation policy (binary variable) |

To assess the mobility changes, we computed the reduction of the mobility metrics with respect to a baseline. The baseline values for each day of the week were obtained by averaging each mobility metrics over the 5 weeks before the outbreak start. The reduction of each mobility metrics on a given day corresponds to the relative difference of the metrics value between the day considered and the baseline day using as a reference value the baseline mobility metrics value.

### 4.5 Regression model

To investigate possible relationships between the reduction in mobility and socioeconomic factors, we performed a regression analysis respectively on 94 provinces (i.e. provinces with a users’ sample population greater than 100) and on the districts of Milan, Rome and Turin. Both for the cities and the provinces, we first performed a variable selection through a Lasso regression [39] for which the objective function is given by:

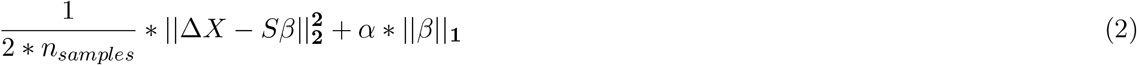

where *n*_*samples*_ is the number of data points, Δ*X* is a vector containing the reductions in mobility (either in radius of gyration or in degree) for each province (resp. urban area) and *S* is a matrix containing the values of the socioeconomic and epidemiological covariates for each province (resp. urban area). The vector *β* is a vector of the regression coefficient values. The covariates selected through the Lasso regression correspond to those obtained with the best model according to cross-validation. We considered penalty parameters *α* values in the following interval [10^*−*3^, 10]. For the provinces, we respectively used 10 and 3 folds for the cross-validation for the provinces and the cities. Once we selected the variables for each day of the three weeks considered, we run an ordinary least squared regression using these variables. The analysis have been done using the Python library *scikit-learn* for the Lasso regression and using the Python module *statsmodels* for the linear regression.

## Data Availability

Anonymised mobility data used in this study, aggregated at the level of Italian provinces, are available at: https://doi.org/10.6084/m9.figshare.c.5029499.v2.
This research also relies on data that is publicly available from the Italian National Institute of Statistics. Data sources are reported in the main text.

## 5 Data availability

Anonymised mobility data used in this study, aggregated at the level of Italian provinces, are available at: https://doi.org/10.6084/m9.figshare.c.5029499.v2. This research also relies on data that is publicly available from the Italian National Institute of Statistics. Data sources are reported in the main text.

## Acknowledgements

EP, LG, CC, MT gratefully acknowledge the support of the Lagrange Project funded by CRT Foundation. PB acknowledges support from Intesa Sanpaolo Innovation Center. This work has received funding from the European Union’s Horizon 2020 research and innovation programme - project EpiPose (No 101003688). The funders had no role in study design, data collection and analysis, decision to publish, or preparation of the manuscript.

## 7 Author contributions statement

LG, PB, MT designed the the study. LG, PB collected and analyzed the data. LG, PB, MT discussed and interpreted the results. EP wrote the code to generate and analyze the mobility metrics. BL, FP, collected the mobility data. LG, PB, MT wrote the manuscript. All authors reviewed and approved the final version of the manuscript.

## 9 Supplementary Information

**Figure S1:**
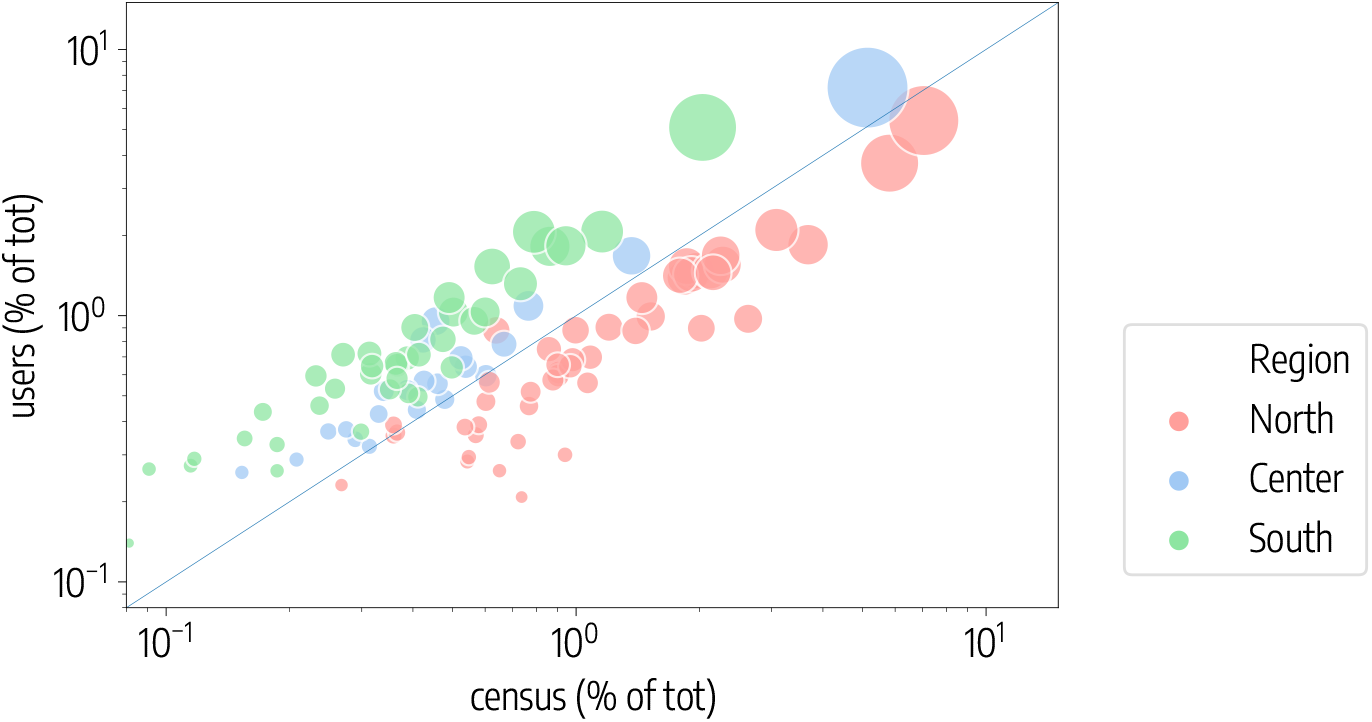
Scatterplot of the percentage of users assigned to each province against the resident population reported by the Italian census, over the total.

**Figure S2:**
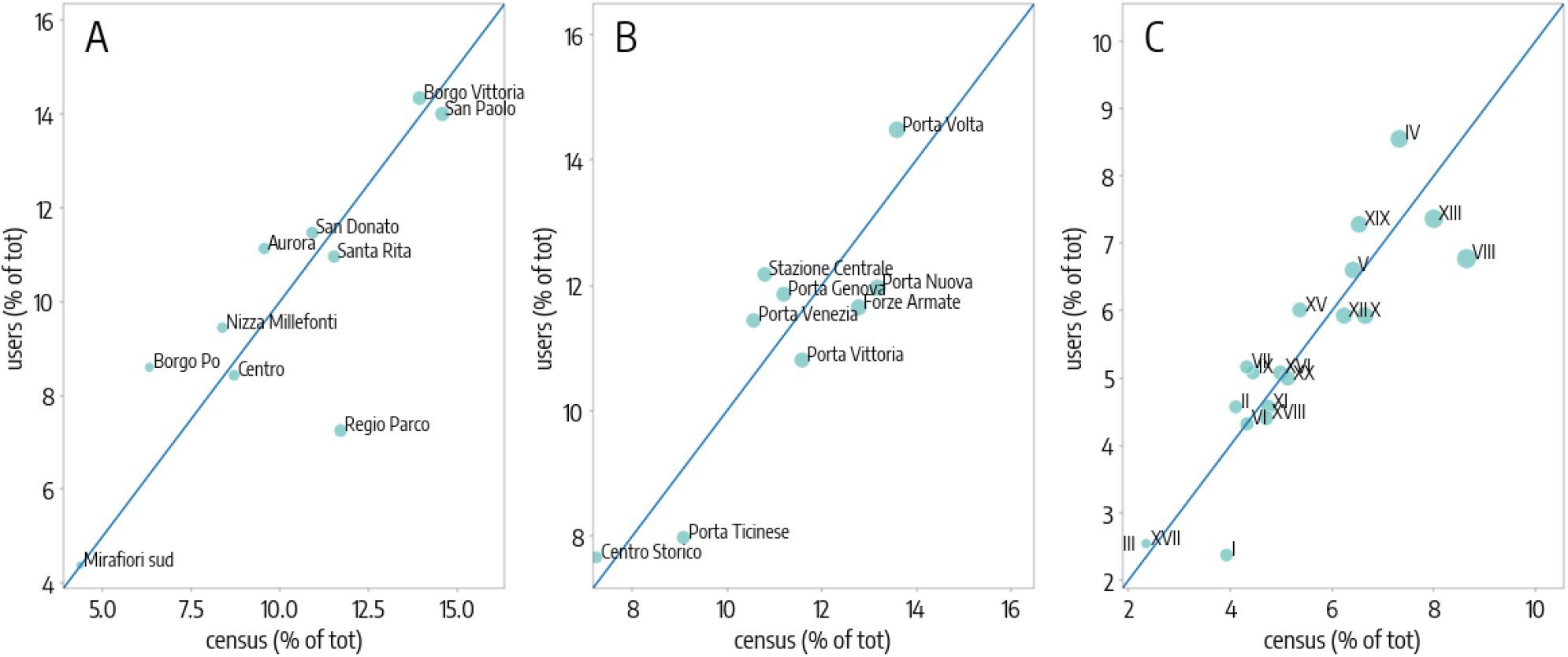
Scatterplot of the percentage of users assigned to each city district against the resident population reported by the Italian census, over the total for A) Turin, B) Milan, C) Rome.

**Table S1:** Socioeconomic determinants of mobility reduction at the province level. The table gives the results of the regression model used to study the relationship between the socioeconomic metrics and the reduction in radius of gyration: we show the r-squared values and the coefficient values with the 95% confidence interval for socioeconomic variables selected by the Lasso regularization, on each day [from Monday to Sunday] during pre-lockdown.

|  | 02 Mar | 03 Mar | 04 Mar | 05 Mar | 06 Mar | 07 Mar | 08 Mar |
| --- | --- | --- | --- | --- | --- | --- | --- |
| r-squared | < 0.1 | < 0.1 | < 0.1 | < 0.1 | < 0.1 | 0.34 | < 0.1 |
| large gatherings ban | 2.1<br>[-1.2, 5.3] | -<br>- | -<br>- | -<br>- | -<br>- | -<br>- | -<br>- |
| attack rate | -<br>- | -<br>- | 1.3<br>[-1.8, 4.4] | -<br>- | -<br>- | 1.5<br>[-2.2, 5.2] | -<br>- |
| unemployment | -<br>- | -<br>- | -<br>- | 3.8<br>[-0.2, 7.8] | -<br>- | 9.1<br>[1.8, 16.4] | 8.0<br>[2.6, 13.4] |
| bar/restaurant SD order | -<br>- | -<br>- | -<br>- | -<br>- | -<br>- | 0.7<br>[-3.6, 4.9] | -<br>- |
| old age index | -<br>- | -<br>- | -<br>- | -<br>- | -<br>- | 1.4<br>[-2.3, 5.1] | -2.2<br>[-6.3, 2.0] |
| population density | -<br>- | -<br>- | -<br>- | -<br>- | -<br>- | 2.6<br>[-1.4, 6.5] | -<br>- |
| high education | -<br>- | -<br>- | -<br>- | -<br>- | -<br>- | 2.2<br>[-1.5, 5.8] | -<br>- |
| commuters | -<br>- | -<br>- | -<br>- | -<br>- | -<br>- | -1.6<br>[-6.2, 3.0] | -<br>- |
| agriculture | -<br>- | -<br>- | -<br>- | -<br>- | -<br>- | -0.5<br>[-5.4, 4.5] | -1.1<br>[-5.6, 3.3] |
| industry | -<br>- | -<br>- | -<br>- | -<br>- | -<br>- | 10.9<br>[5.7, 16.0] | 5.7<br>[0.5, 10.9] |
| income | -<br>- | -<br>- | -<br>- | -<br>- | -<br>- | -8.5<br>[-16.1, -0.9] | -<br>- |
| services | -<br>- | -<br>- | -<br>- | -<br>- | -<br>- | -<br>- | -<br>- |

**Table S2:**
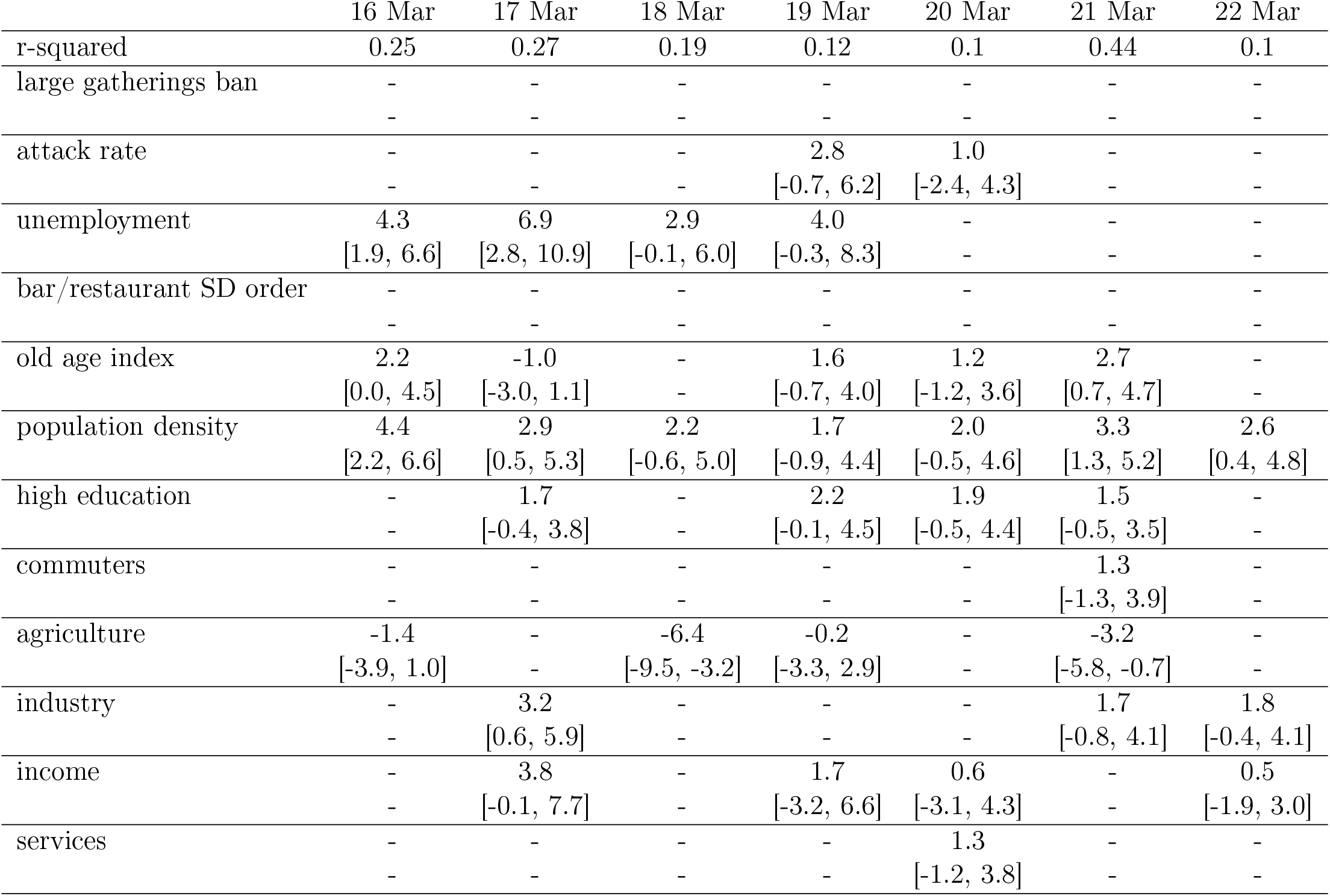
Socioeconomic determinants of mobility reduction at the province level. The table gives the results of the regression model used to study the relationship between the socioeconomic metrics and the reduction in radius of gyration: we show the r-squared values and the coefficient values with the 95 % confidence interval for socioeconomic variables selected by the Lasso regularization, on each day [from Monday to Sunday] during lockdown.

**Table S3:**
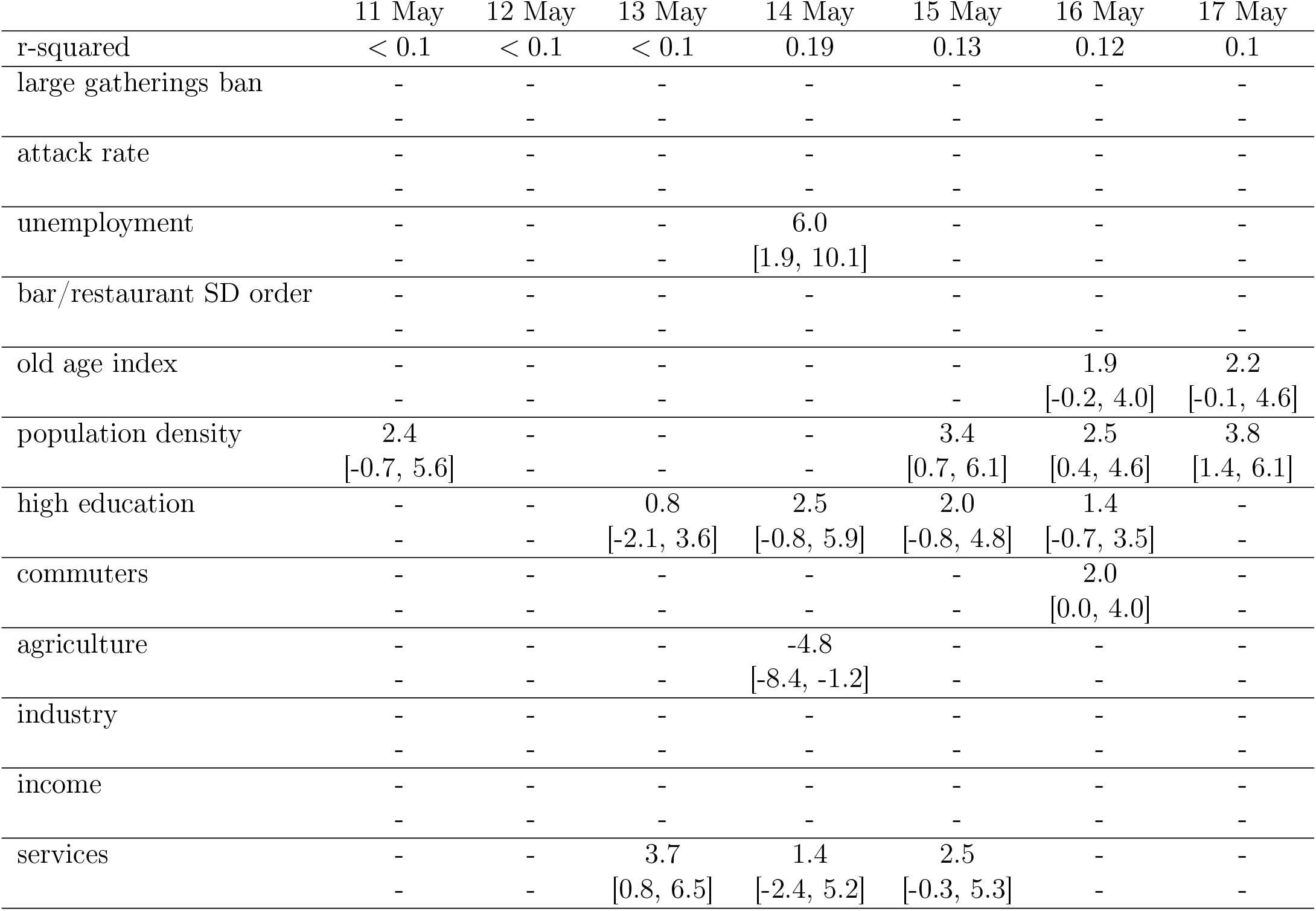
Socioeconomic determinants of mobility reduction at the province level. The table gives the results of the regression model used to study the relationship between the socioeconomic metrics and the reduction in radius of gyration: we show the r-squared values and the coefficient values with the 95 % confidence interval for socioeconomic variables selected by the Lasso regularization, on each day [from Monday to Sunday] during phase 2.

**Table S4:**
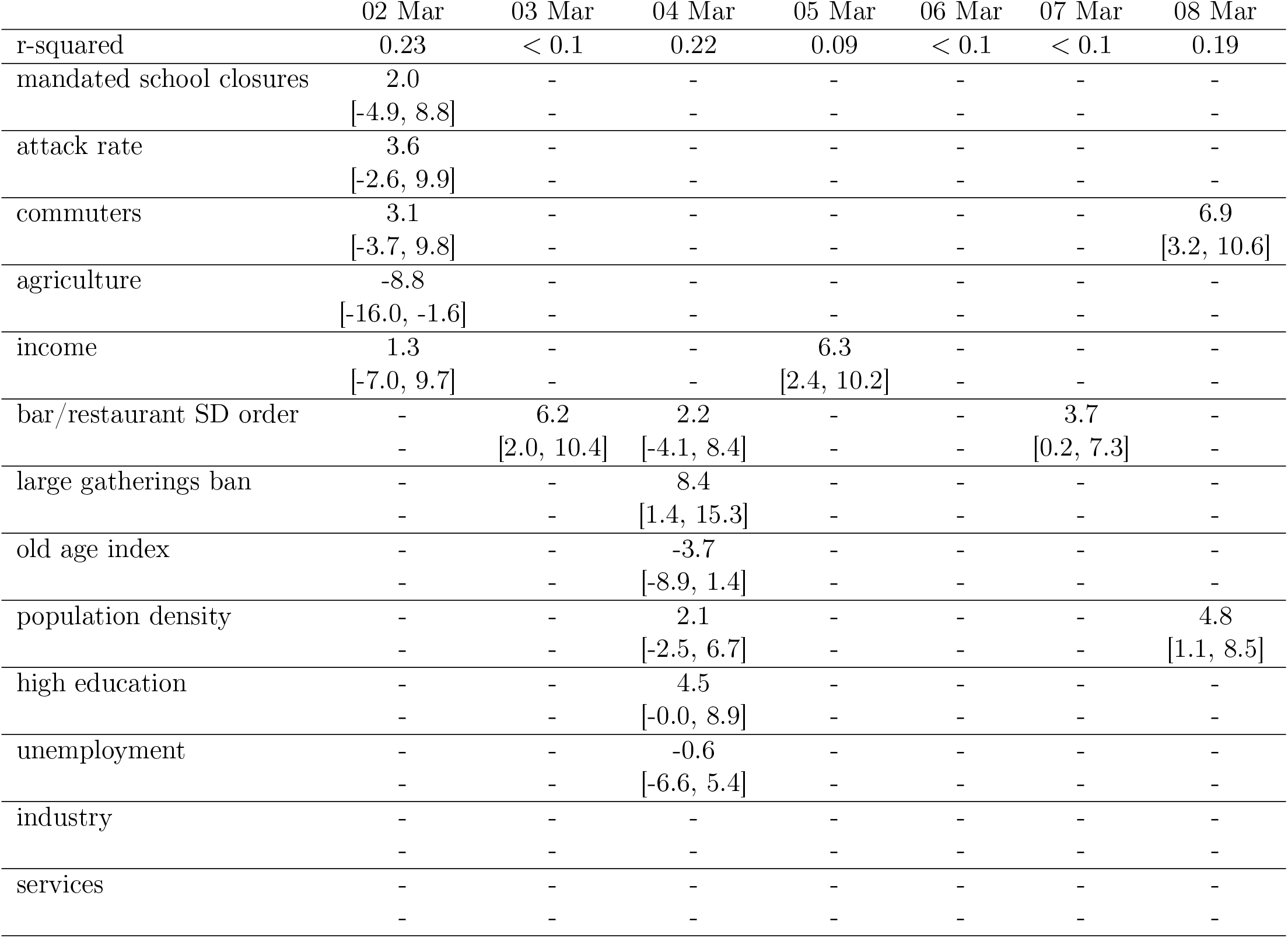
Socioeconomic determinants of mobility reduction at the province level. The table gives the results of the regression model used to study the relationship between the socioeconomic metrics and the reduction in degree: we show the r-squared values and the coefficient values with the 95 %confidence interval for socioeconomic variables selected by the Lasso regularization, on each day [from Monday to Sunday] during pre-lockdown.

**Table S5:**
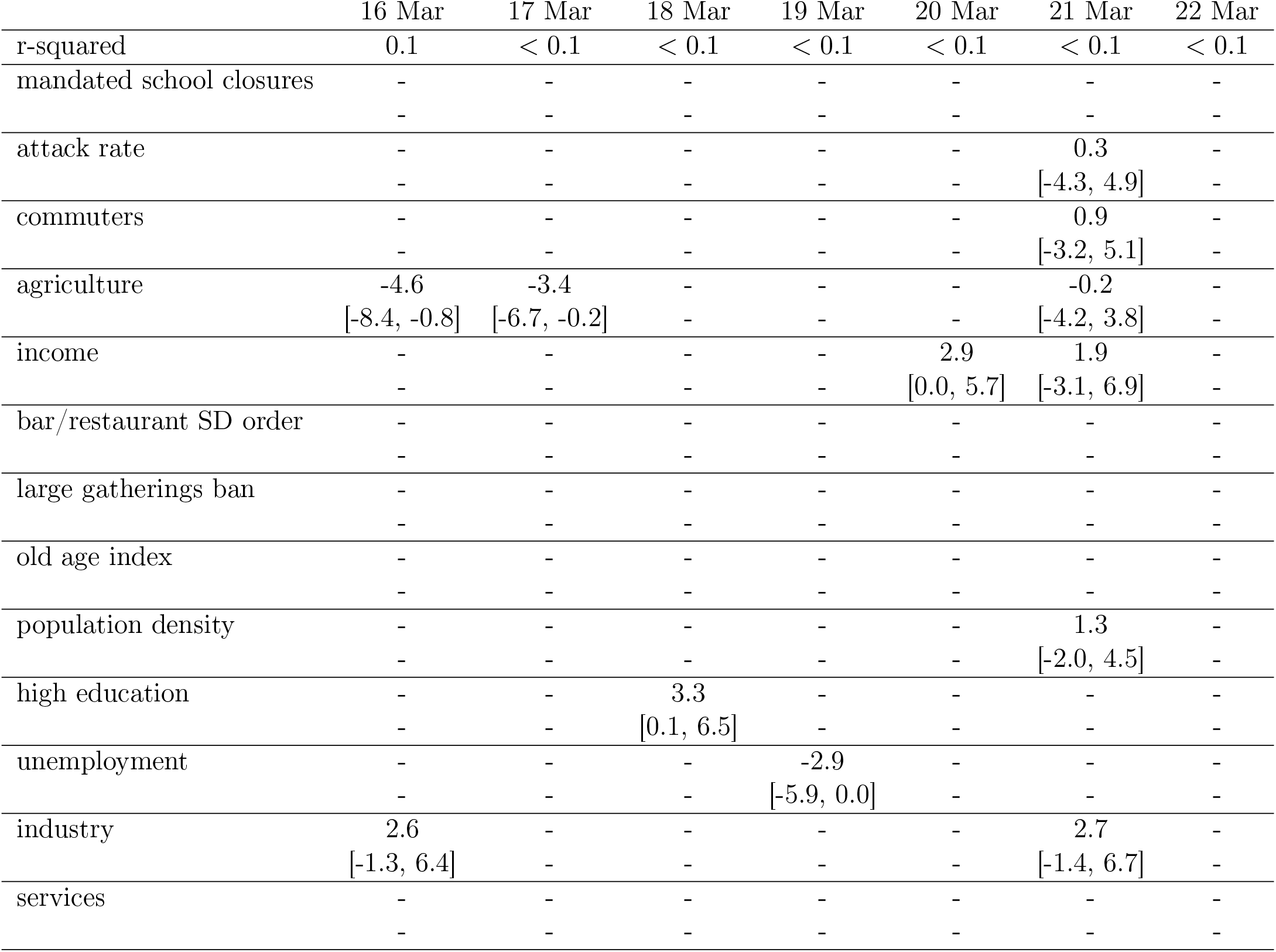
Socioeconomic determinants of mobility reduction at the province level. The table gives the results of the regression model used to study the relationship between the socioeconomic metrics and the reduction in radius of gyration: we show the r-squared values and the coefficient values with the 95 % confidence interval for socioeconomic variables selected by the Lasso regularization, on each day [from Monday to Sunday] during lockdown.

**Table S6:**
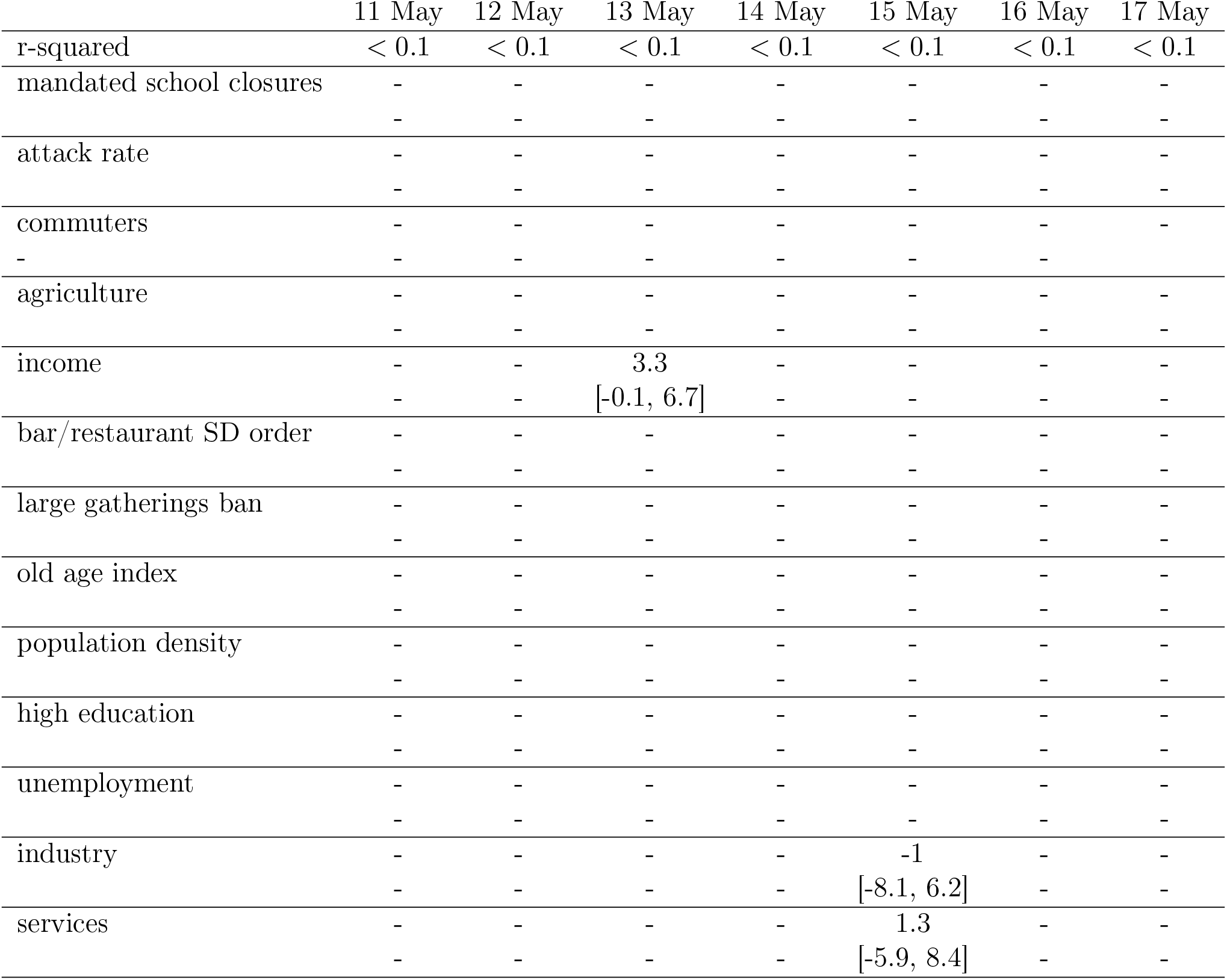
Socioeconomic determinants of mobility reduction at the province level. The table gives the results of the regression model used to study the relationship between the socioeconomic metrics and the reduction in degree: we show the r-squared values and the coefficient values with the 95 %confidence interval for socioeconomic variables selected by the Lasso regularization, on each day [from Monday to Sunday] during phase 2.

**Table S7:**
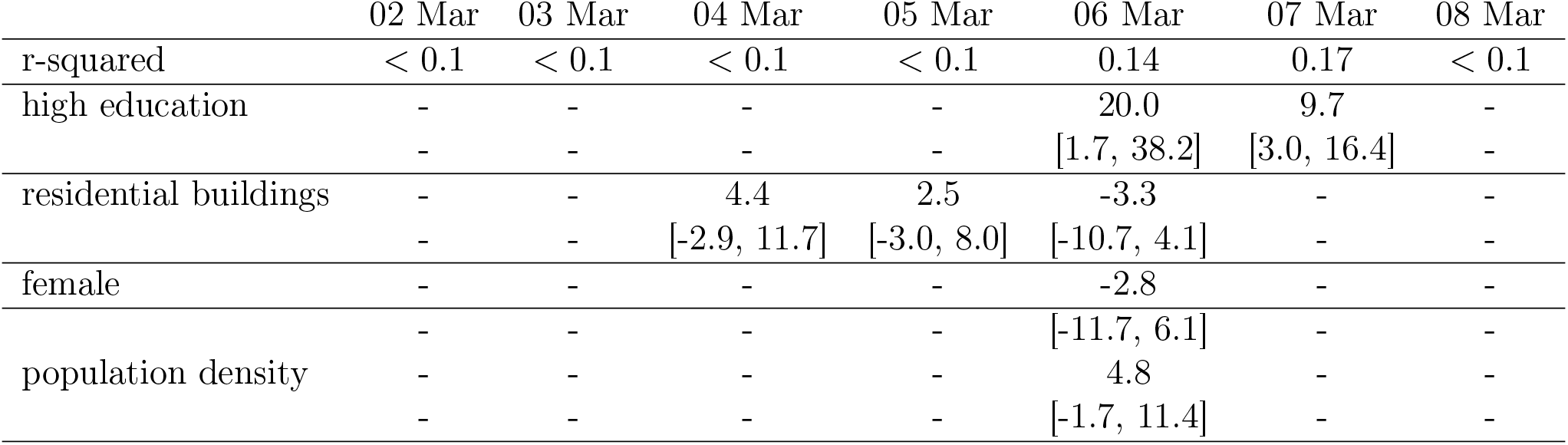
Socioeconomic determinants of mobility reduction at the city district level. The table gives the results of the regression model used to study the relationship between the socioeconomic metrics and the reduction in degree: we show the r-squared values and the coefficient values with the 95 %confidence interval for socioeconomic variables selected by the Lasso regularization, on each day [from Monday to Sunday] during pre-lockdown.

**Table S8:**
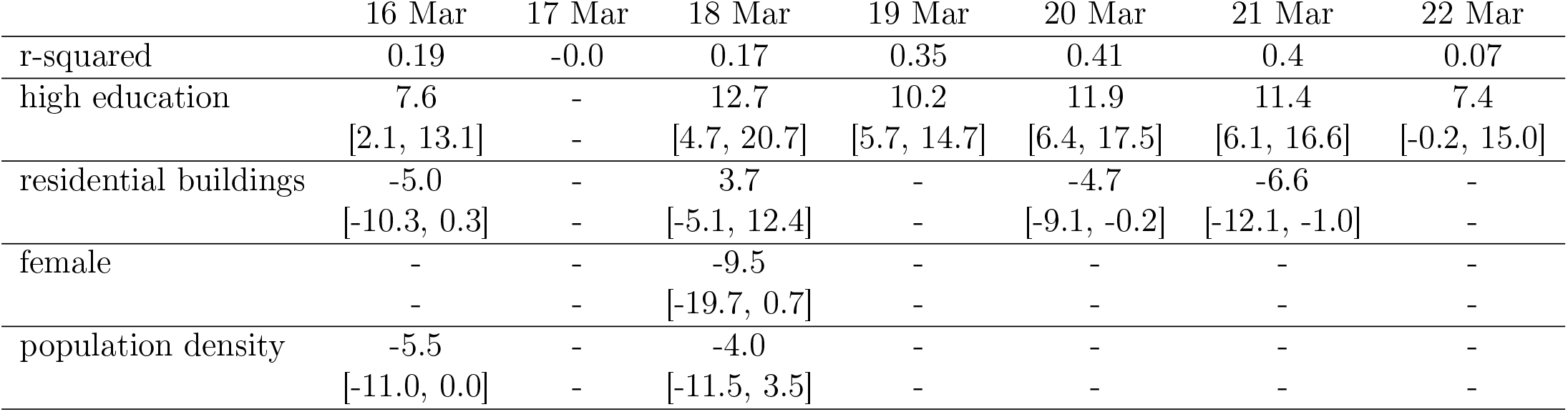
Socioeconomic determinants of mobility reduction at the city district level. The table gives the results of the regression model used to study the relationship between the socioeconomic metrics and the reduction in degree: we show the r-squared values and the coefficient values with the 95 %confidence interval for socioeconomic variables selected by the Lasso regularization, on each day [from Monday to Sunday] during lockdown.

**Table S9:**
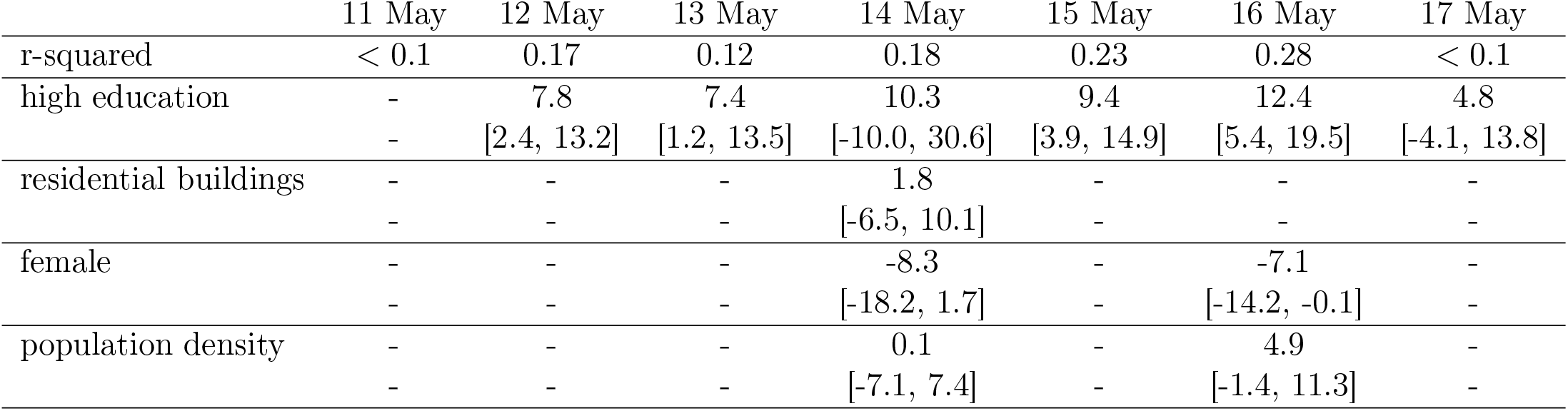
Socioeconomic determinants of mobility reduction at the city district level. The table gives the results of the regression model used to study the relationship between the socioeconomic metrics and the reduction in degree: we show the r-squared values and the coefficient values with the 95 %confidence interval for socioeconomic variables selected by the Lasso regularization, on each day [from Monday to Sunday] during phase 2.

**Figure S3:**
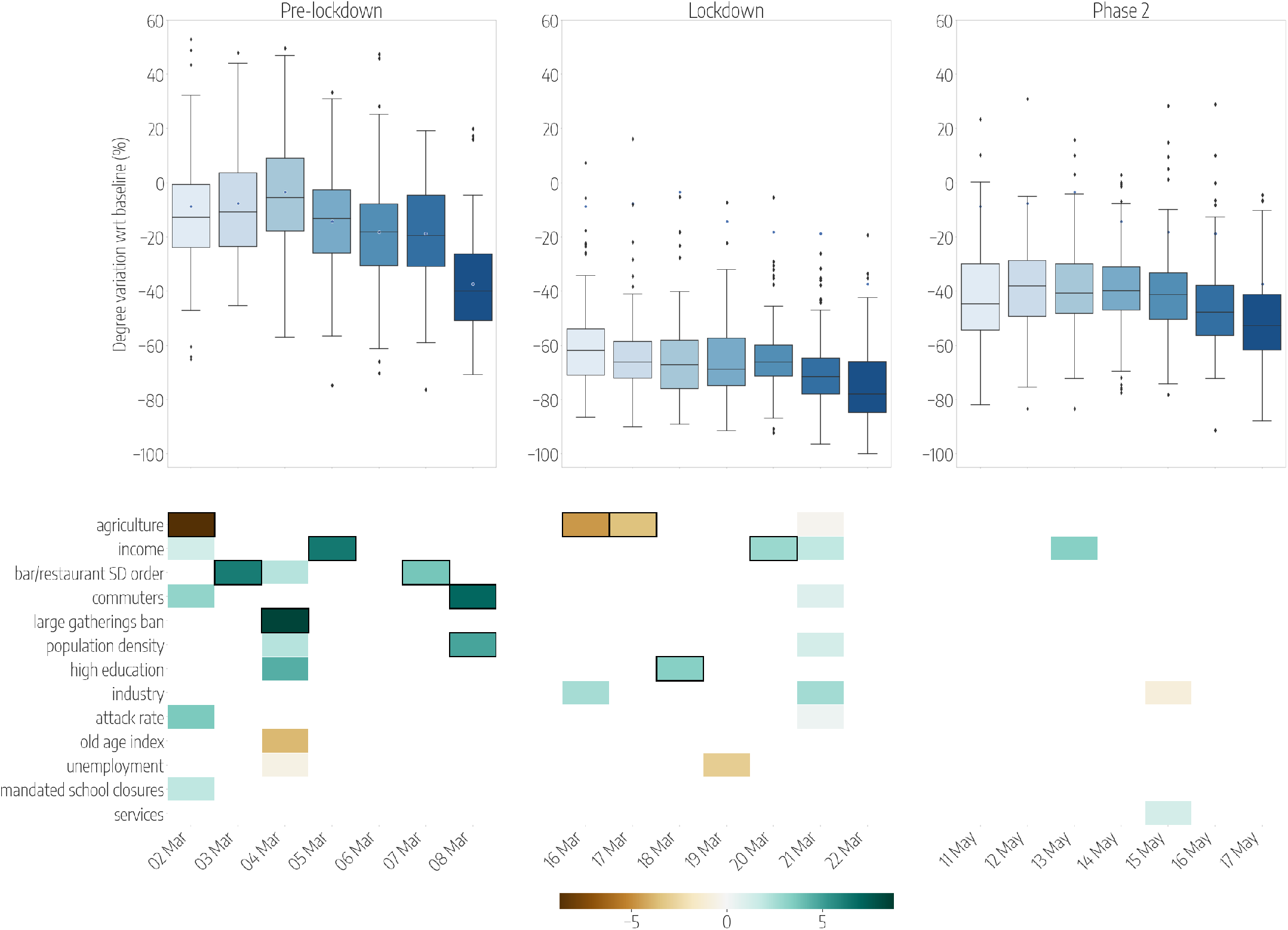
Province-level mobility reductions and their socioeconomic determinants. Top panels show the relative reductions in the median degree with respect to the baseline, each day (from Monday to Sunday) in 3 different weeks of the periods under study: the Pre-lockdown (left), the Lockdown (center) and the Phase 2 (right). Boxplots describe the distributions of the reductions by province. Bottom panels show the results of the regression model, by week, displaying the socioeconomic variables that are selected by the model to predict the corresponding variations in mobility, on each day. Coloured boxes correspond to those covariates that are selected by the model, and the colour codes the associated coefficient in the regression, from negative (brown) to positive (blue). Black solid squares highlight the covariates that are statistically significant at *p <* 0.05.

**Figure S4:**
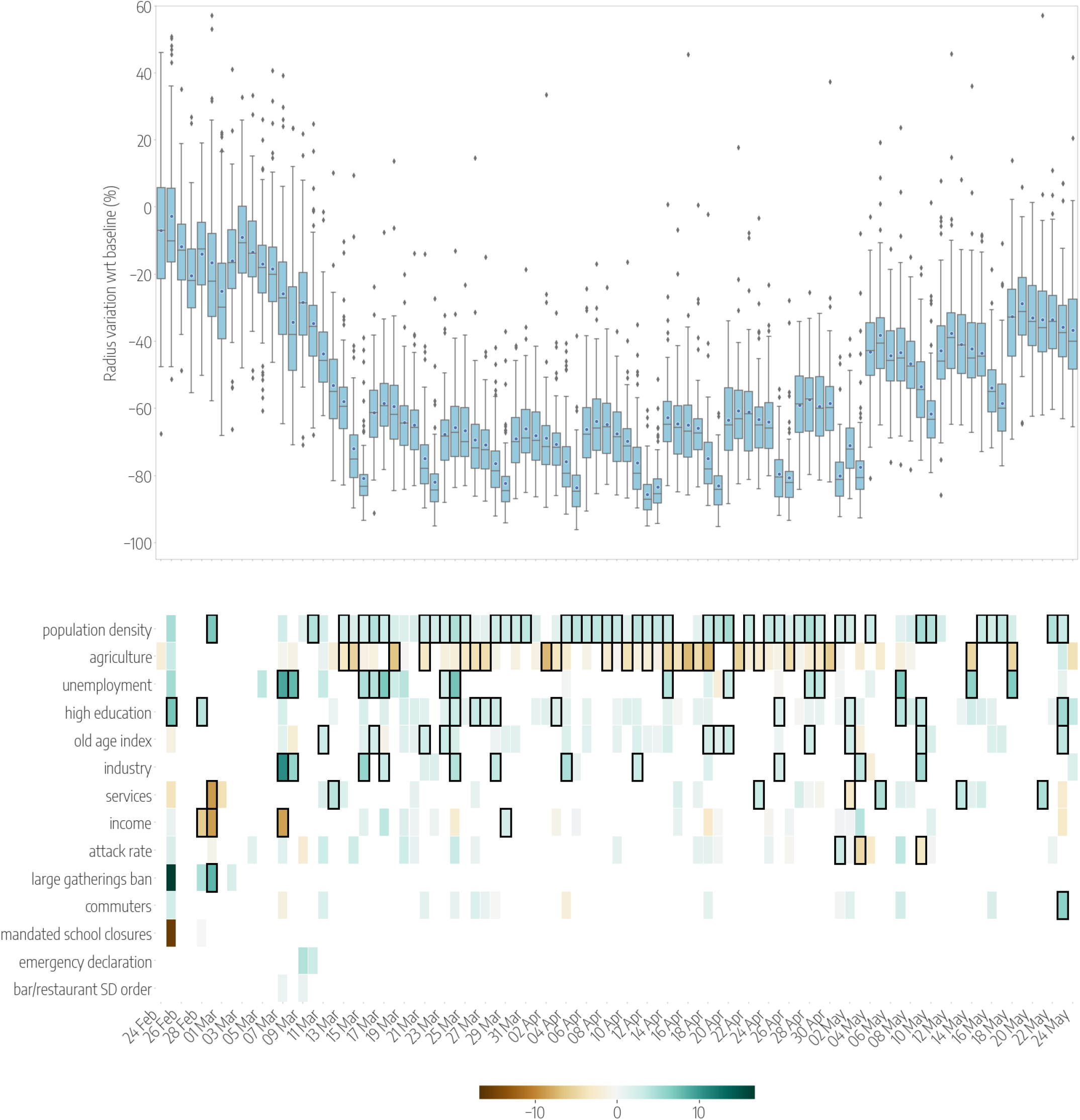
Province-level mobility reductions and their socioeconomic determinants. Top panels show the relative reductions in the mean radius of gyration with respect to the baseline during the whole timeline of the dataset. Boxplots describe the distributions of the reductions by province. Bottom panels show the results of the regression model, displaying the socioeconomic variables that are selected by the model to predict the corresponding variations in mobility, on each day. Coloured boxes correspond to those covariates that are selected by the model, and the color codes the associated coefficient in the regression, from negative (brown) to positive (blue). Black solid squares highlight the covariates that are statistically significant at *p <* 0.05.

**Figure S5:**
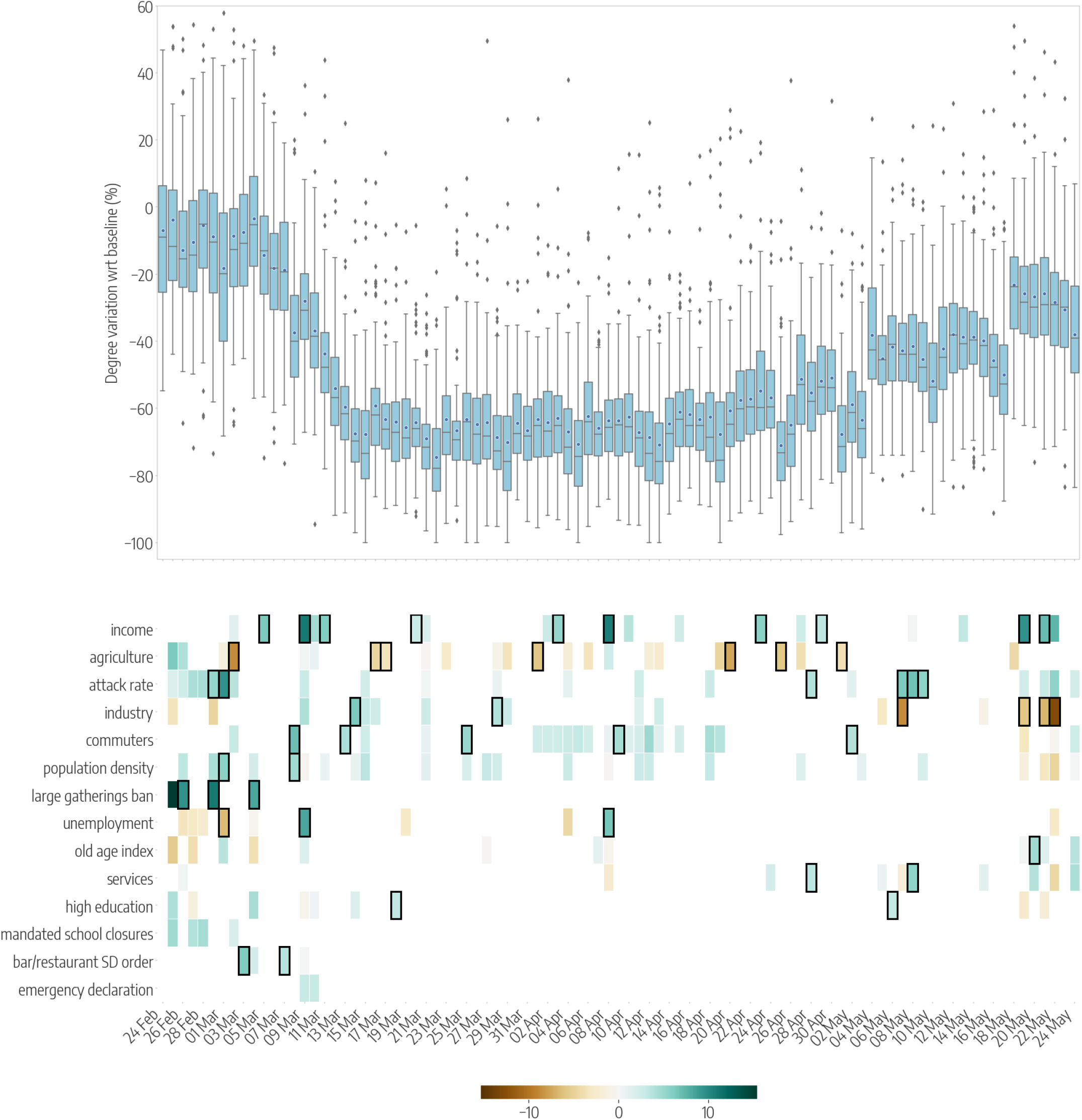
Province-level mobility reductions and their socioeconomic determinants. Top panels show the relative reductions in the mean degree with respect to the baseline during the whole timeline of the dataset. Boxplots describe the distributions of the reductions by province. Bottom panels show the results of the regression model, displaying the socioeconomic variables that are selected by the model to predict the corresponding variations in mobility, on each day. Coloured boxes correspond to those covariates that are selected by the model, and the color codes the associated coefficient in the regression, from negative (brown) to positive (blue). Black solid squares highlight the covariates that are statistically significant at *p <* 0.05.

**Figure S6:**
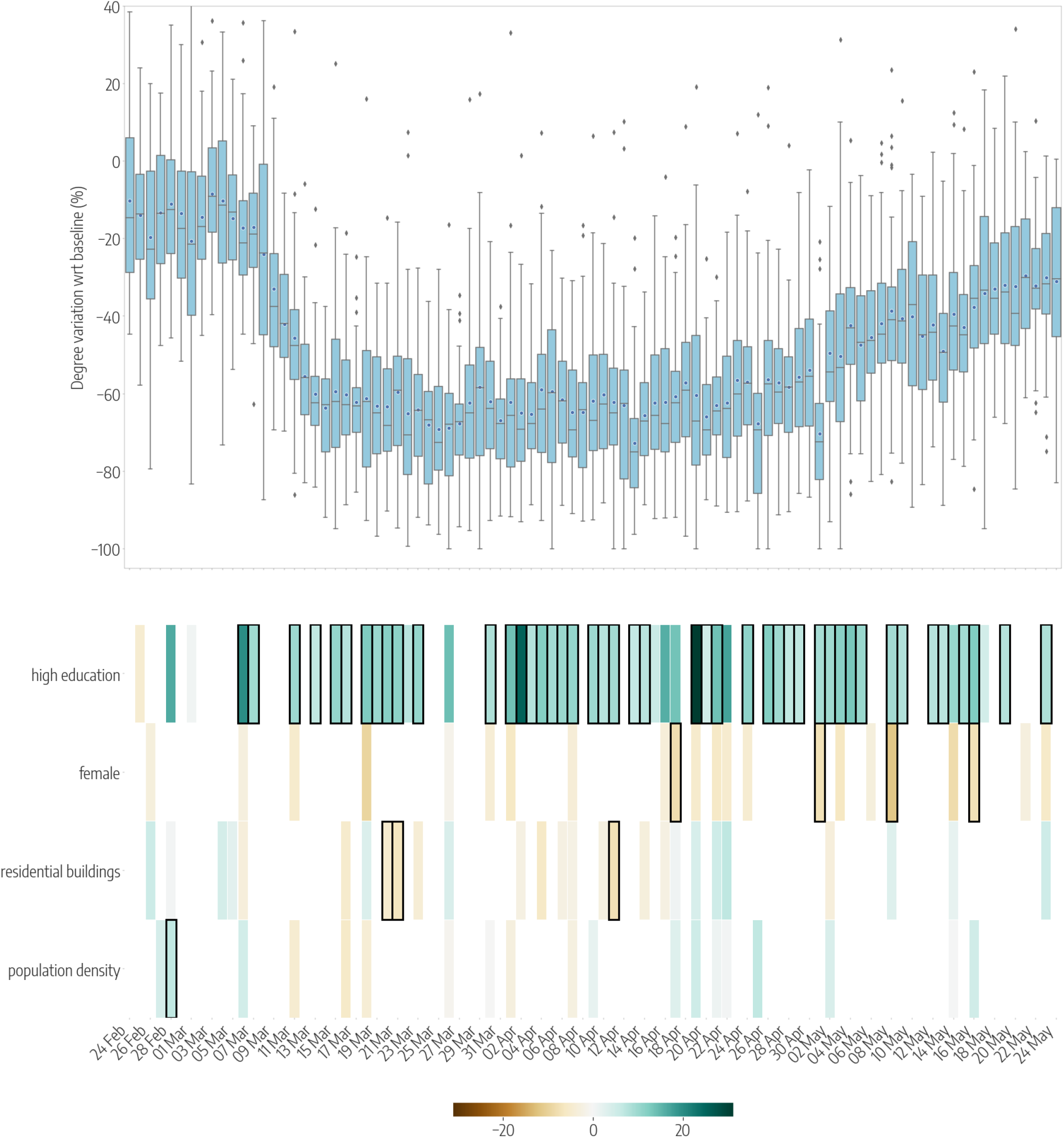
City district level mobility reductions and their socioeconomic determinants. Top panels show the relative reductions in the mean degree with respect to the baseline during the whole timeline of the dataset. Boxplots describe the distributions of the reductions by city distric. Bottom panels show the results of the regression model, displaying the socioeconomic variables that are selected by the model to predict the corresponding variations in mobility, on each day. Coloured boxes correspond to those covariates that are selected by the model, and the color codes the associated coefficient in the regression, from negative (brown) to positive (blue). Black solid squares highlight the covariates that are statistically significant at *p <* 0.05.

